# Persistent Orofacial Pain Attendances at General Medical Practitioners

**DOI:** 10.1101/2022.05.30.22275776

**Authors:** CC Currie, J Palmer, SJ Stone, P Brocklehurst, VR Aggarwal, PJ Dorman, MS Pearce, J Durham

## Abstract

Patients with persistent orofacial pain (OFP) can go through complex care pathways to receive a diagnosis and management, which can negatively impact their pain. This study aimed to describe 44-year trends in attendances at Welsh medical practices for persistent OFP and establish the number of attendances per patient and referrals associated with OFP and factors which may predict whether a patient is referred. A retrospective observational study was completed using the nationwide Secure Anonymised Information Linkage Databank of visits to general medical practices in Wales (UK). Orofacial and migraine Read codes were extracted between 1974 and 2017. Data were analysed using descriptive statistics, univariate, and multivariable logistic regression. Over the 44-year period there were 468,827 persistent OFP and migraine Read codes, accounting for 468,137 patient attendances, or 301,832 patients. The overall attendance rate was 4.22 attendances per 1000 patient-years (95% CI 4.21-4.23). The attendance rate increased over the study period. Almost one-third of patients (n=92,192, 30.54%) attended more than once over the study period and 15.83% attended more than once within a 12-month period. There were 20,103 referral Read codes which were associated with 8,183 patients, with over half these patients being referred more than once. Odds of receiving a referral were highest in females (OR 1.23; 95% CI 1.17-1.29), in those living in rural locations (OR 1.17; 95% CI 1.12-1.22) and in the least deprived quintile (OR 1.39; 95% CI 1.29-1.48). Odds also increased with increasing age (OR 1.03; 95% CI 1.03-1.03). The increasing attendance may be explained by the increasing incidence of persistent OFP within the population. Current care pathways when these patients do seek care from their GMP could be improved. Referrals could be encouraged at an earlier point and made to the most appropriate place to streamline care pathways which could result in improved patient outcomes.

## Introduction

Orofacial pain (OFP) is one of the most common causes of persistent (also previously known as chronic) pain (Breivik et al. 2006) affecting around 7% of the UK population (Aggarwal et al. 2006). Persistent OFP can encompass a number of conditions/disorders including temporomandibular disorders (TMD), persistent idiopathic orofacial pain, burning mouth syndrome (BMS), post-traumatic trigeminal neuropathic pain and trigeminal neuralgia (International Classification of Orofacial Pain 2020). The most common is TMD, a collective term for a group of musculoskeletal conditions involving pain and/or dysfunction in the muscles of mastication, temporomandibular joint and associated structures (Maixner et al. 2011). Migraine can also present in the face and be considered a persistent OFP diagnosis (Headache Classification Subcommitee of the International Headache Society 2013; International Classification of Orofacial Pain 2020) and can also co-exist with other persistent or chronic OFP diagnoses, for example patients with TMD have four to five times the odds of having co-existing migraine (Réus et al. 2022). Persistent OFP also co-occurs with other persistent pain conditions like chronic widespread pain, irritable bowel syndrome and low back pain and may be part of a wider spectrum of pain disorders associated with increased psychosocial co-morbidities (Aggarwal et al. 2006).

Persistent OFP exerts a substantial effect on quality of life (Shueb et al. 2015) and a substantial economic impact (Durham et al. 2016; Breckons et al. 2018). When patients experience OFP they can present to a range of healthcare professionals including general dental practitioners (GDPs) and general medical practitioners (GMPs), being informally referred between the two as well as being referred to multiple secondary care services (Breckons et al. 2017). Evidence suggests that healthcare professionals find persistent OFP difficult to diagnose and manage (Peters et al. 2015), leading to complex care pathways for these patients which negatively impacts on their pain and long term management (Durham et al. 2010; Durham et al. 2021). Although attendances at GMPs for dental problems has been investigated (Anderson et al. 1999; Cope et al. 2016; Currie et al. 2022), the number of patients attending specifically with persistent OFP diagnoses has not.

The aim of this study was to describe 44-year trends in attendances at GMPs for persistent OFP. Specific objectives were to explore the number of attendances and sociodemographic factors of patients attending here, and establish the number of referrals associated with orofacial pain and factors which may predict whether a patient is referred.

## Materials and Methods

The study details have been described in detail elsewhere (Currie et al. 2022). In brief, a retrospective observational study was completed using the General Practitioner (GP) dataset within the Secure Anonymised Information Linkage (SAIL) Databank (Ford et al. 2009). SAIL is a national dataset consisting of anonymised health and administrative datasets from the Welsh population and contains a “GP dataset” with over 40 years of data on Welsh GMP attendances. This gave annual, cross-sectional data on patient attendances for persistent OFP for each of the 44 years. Approval was granted by the Health Information Research Unit Information Governance Review Panel.

Data were identified and extracted by a SAIL analyst. At the time of data extraction, the dataset covered 76.9% of GMP practices. All patient attendances for persistent OFP were included between 1^st^ January 1974 and 31^st^ December 2017. Identification of relevant patient attendances was with dental and orofacial Read codes (version 2) (Appendix Table 1). Read codes are a clinical terminology used in General Medical Practice in the UK based on medical terms and include and cross reference all of the other widely used medical classifications, they are used to code details of multiple demographics, investigations, therapeutics and operative treatments of individual patients (Chisholm 1990). Acute dental pain Read codes were excluded, however non-specific dental Read codes which could encompass symptoms of persistent OFP (e.g., tooth symptoms) were included (Appendix Table 1). Read codes for migraine were included to encompass both migraine as a potential persistent OFP diagnosis, and as a comparator against other diagnoses.

For each Read code identified the following covariates were extracted: patient ID, week of birth, gender, Welsh Index of Multiple Deprivation (WIMD) quintile, Urban/Rural classification, attendance date. WIMD is the official measure of relative deprivation of areas of Wales (Welsh Government 2011), and the Office for National Statistics Urban/Rural classification 2001 (Office for National Statistics 2004) divides areas in urban and rural categories with further subdivisions by sparsity (further details available in the appendix). Patient age was calculated using week of birth and attendance date. For each persistent OFP attendance identified associated referral Read codes were also included. Rate of attendance was calculated as number of attendances over time and converted into attendance rates per 1000 patient-years using the Welsh Demographic Service dataset (supplemental methodological details available in the Appendix).

Data cleaning was undertaken prior to analysis with STATA v15 (Statacorp LP, College Station, TX, USA) within the SAIL portal. To protect patients’ confidentiality, counts less than five were not exported from the portal, therefore Read codes were grouped into larger diagnostic groups (Appendix Table 1). Where regrouping was not possible counts were denoted as “<5”, and the total number for that variable adjusted. Read codes relating to non-specific dental diagnoses/symptoms which could be suggestive of OFP (Appendix Table 1) were grouped to form a “non-specific dental diagnosis” group. To examine predictors of being referred univariate and multivariable logistic regression modelling was performed. The binary response variable was whether a patient was referred or not. Explanatory variables were gender, age, WIMD, urban/rural, and potential confounders and interactions between age, gender, WIMD and urban/rural were assessed. Regression modelling was repeated with adjustments for any potential confounders and included in the final model where a larger than 10% change was observed.

## Results

Over the period studied, there were 468,827 persistent OFP Read codes, accounting for 468,137 patient attendances, or 301,832 patients. The overall attendance rate was 4.22 attendances per 1000 patient-years (95% CI 4.21-4.23), this reduced to 1.53 attendances per 1000 patient-years (95% CI 1.52-1.54) when migraine was excluded. Patients most commonly attended with migraine, followed by non-specific dental diagnoses and TMD. The breakdown by diagnosis is given in Table 1. 5,508 (1.82%) patients had a diagnosis of both migraine and TMD over the time period studied.

**Table 1:**
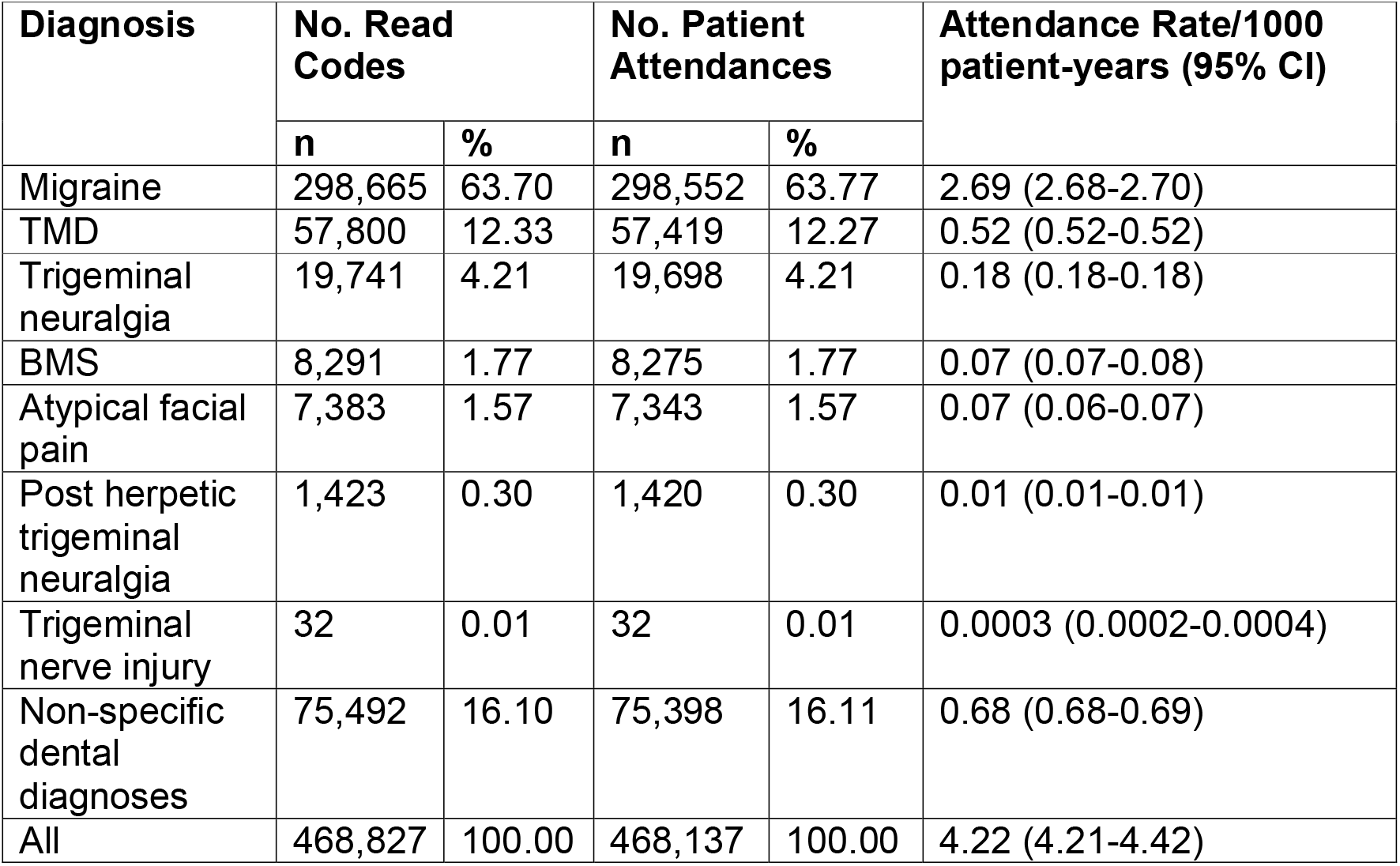
Breakdown of number of attendances by diagnosis during the 44-year study period 1974-2017.

| Diagnosis | No. Read Codes |  | No. Patient Attendances |  | Attendance Rate/1000 patient-years (95% CI) |
| --- | --- | --- | --- | --- | --- |
|  | n | % | n | % |  |
| Migraine | 298,665 | 63.70 | 298,552 | 63.77 | 2.69 (2.68-2.70) |
| TMD | 57,800 | 12.33 | 57,419 | 12.27 | 0.52 (0.52-0.52) |
| Trigeminal neuralgia | 19,741 | 4.21 | 19,698 | 4.21 | 0.18 (0.18-0.18) |
| BMS | 8,291 | 1.77 | 8,275 | 1.77 | 0.07 (0.07-0.08) |
| Atypical facial pain | 7,383 | 1.57 | 7,343 | 1.57 | 0.07 (0.06-0.07) |
| Post herpetic trigeminal neuralgia | 1,423 | 0.30 | 1,420 | 0.30 | 0.01 (0.01-0.01) |
| Trigeminal nerve injury | 32 | 0.01 | 32 | 0.01 | 0.0003 (0.0002-0.0004) |
| Non-specific dental diagnoses | 75,492 | 16.10 | 75,398 | 16.11 | 0.68 (0.68-0.69) |
| All | 468,827 | 100.00 | 468,137 | 100.00 | 4.22 (4.21-4.42) |

Patient attendances for persistent OFP increased from 1988 to 2006 and then remained relatively stable over the remaining study period (Figure 1). Migraine was consistently the most common diagnosis, with all diagnoses showing an increase in attendance rate over the study period except for non-specific dental diagnoses which initially increased then declined following 2012 (Figure 2).

**Figure 1:**
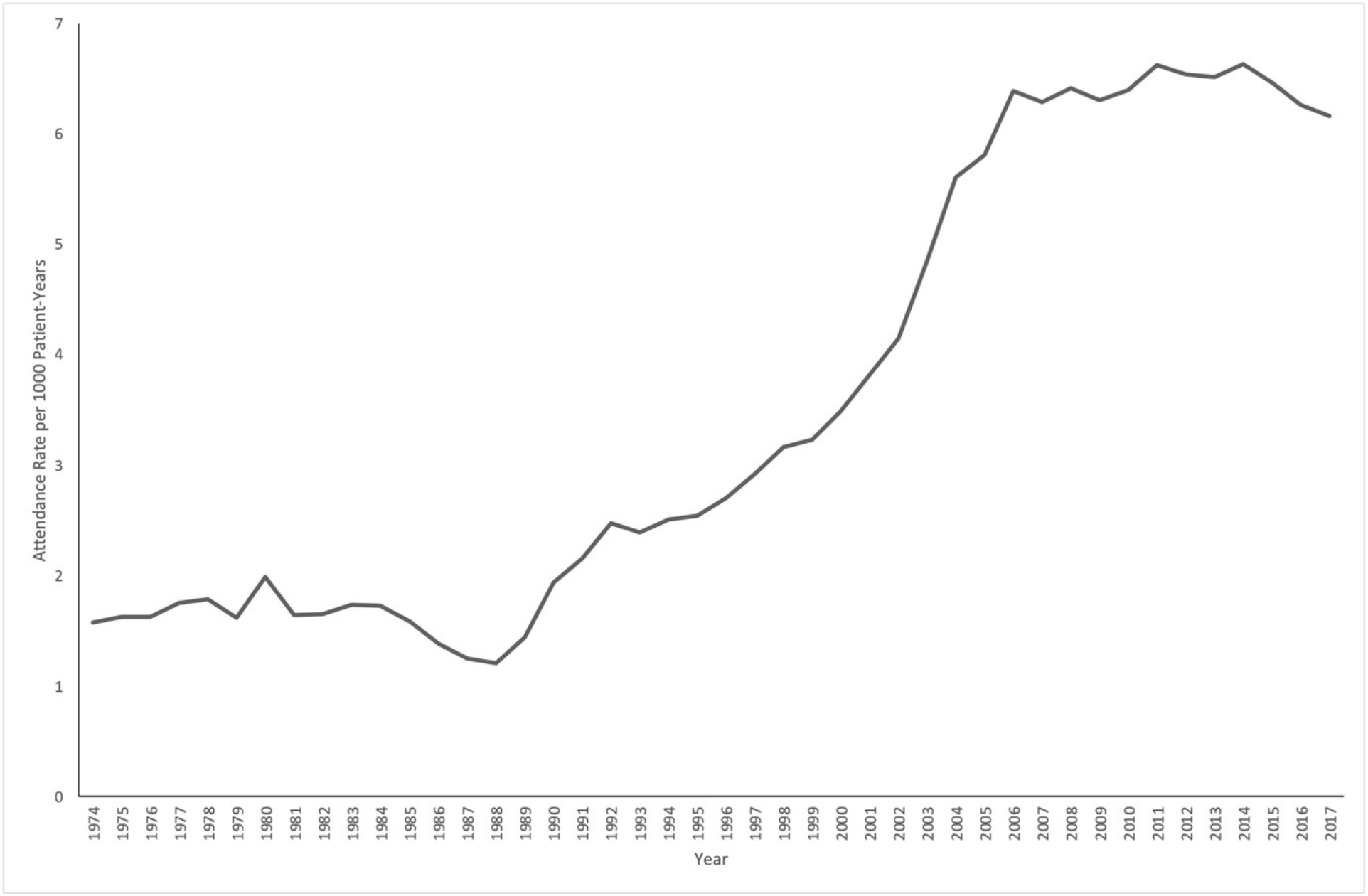
Attendance rate for all patients with persistent OFP diagnoses over the 44-year study period.

**Figure 2:**
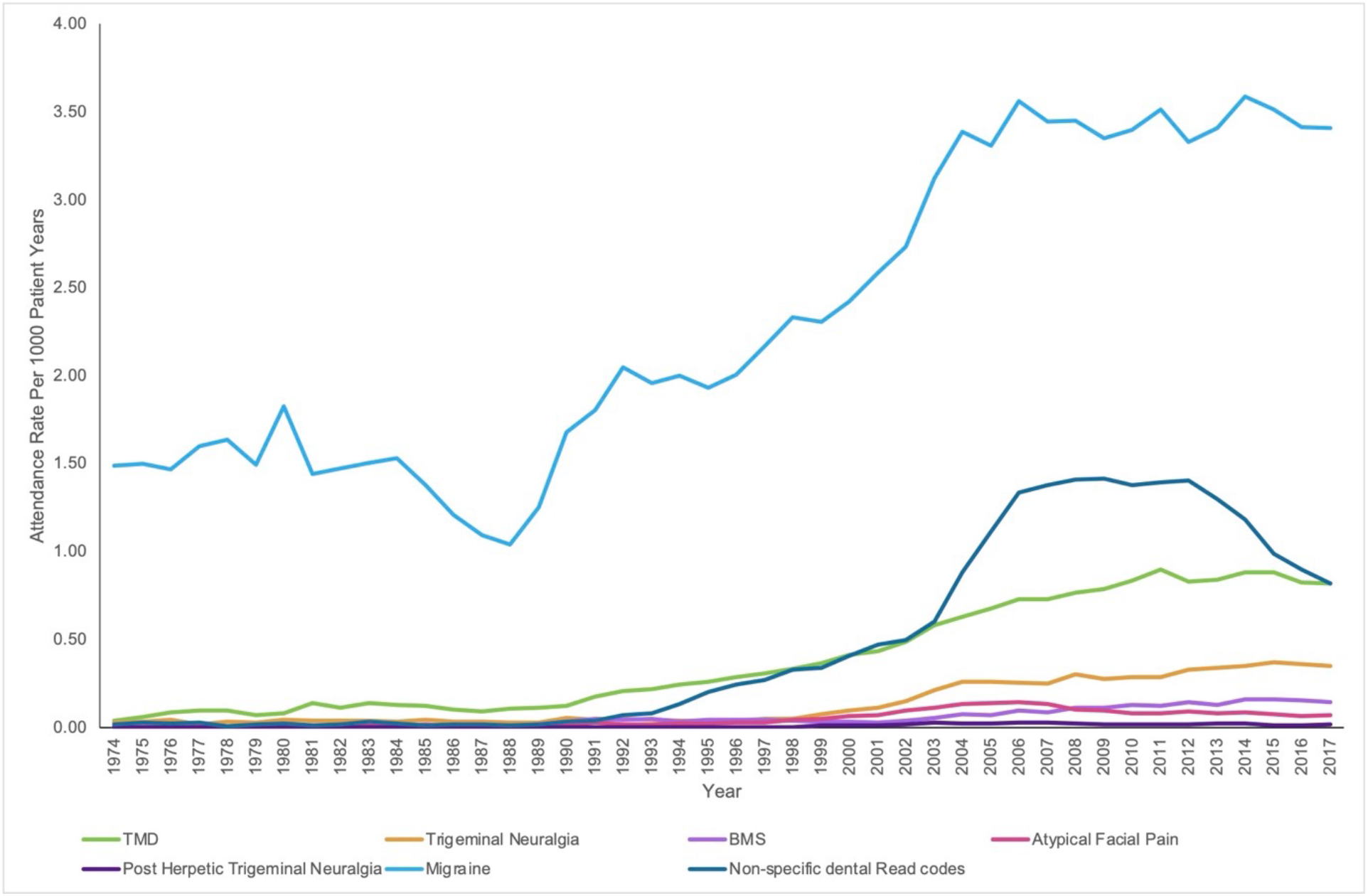
Attendance rate over the 44-year study period by OFP diagnosis.

Detailed patient demographics are given in Appendix Tables 2 and 3. Patients tended to be female (71.66%) and aged 20-29 years (20.58%). Patients were more commonly from urban areas (65.92%) and relatively equally distributed between WIMD quintiles (X^2^ (4df, n=468,137)=32.39, p=0.996).

Almost one-third of patients (n=92,192, 30.54%) attended more than once over the study period. 47,769 patients (15.83%) attended more than once within a 12-month period. The breakdown of number of attendances over the entire study period, and within 12-months are shown in Table 2. The number of attendances for patients with a diagnosis of both migraine and TMD is given in appendix Table 4.

**Table 2:**
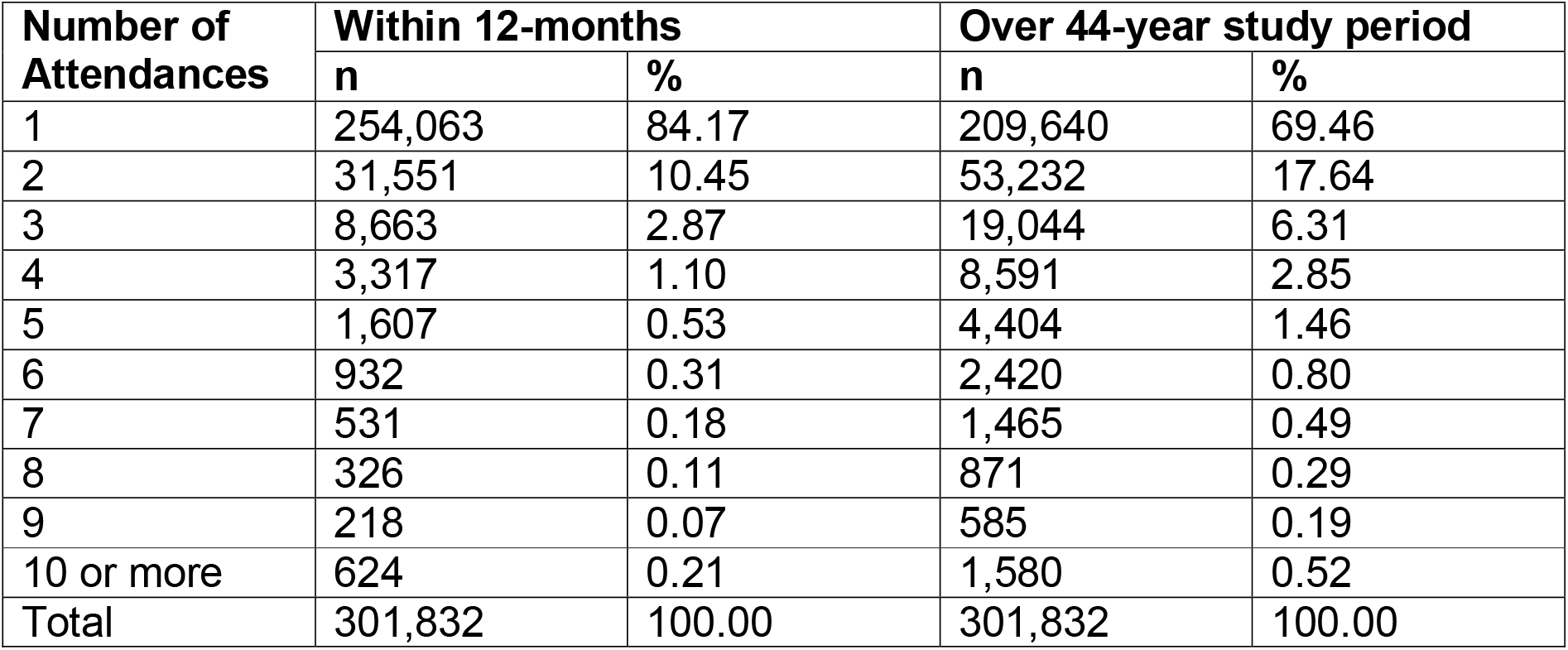
Number of attendances for persistent OFP within 12-months and over the 44-year study period.

| Number of Attendances | Within 12-months |  | Over 44-year study period |  |
| --- | --- | --- | --- | --- |
|  | n | % | n | % |
| 1 | 254,063 | 84.17 | 209,640 | 69.46 |
| 2 | 31,551 | 10.45 | 53,232 | 17.64 |
| 3 | 8,663 | 2.87 | 19,044 | 6.31 |
| 4 | 3,317 | 1.10 | 8,591 | 2.85 |
| 5 | 1,607 | 0.53 | 4,404 | 1.46 |
| 6 | 932 | 0.31 | 2,420 | 0.80 |
| 7 | 531 | 0.18 | 1,465 | 0.49 |
| 8 | 326 | 0.11 | 871 | 0.29 |
| 9 | 218 | 0.07 | 585 | 0.19 |
| 10 or more | 624 | 0.21 | 1,580 | 0.52 |
| Total | 301,832 | 100.00 | 301,832 | 100.00 |

There were 20,103 referral Read codes associated with persistent OFP and migraine diagnostic Read codes. These were associated with 8,183 patients, with over half these patients being referred more than once (Table 3). Referral locations are given in Appendix Table 5 and included a range of healthcare professionals across both National Health Service (NHS) and private providers, as well as referral pathways for suspected head and neck cancer. The number of referrals by diagnosis is given in Appendix Tables 6 and 7.

**Table 3:** Number of times patients were referred over the 44-year study period.

| Number of Times Referred | Number of Patients | Percentage |
| --- | --- | --- |
| 1 | 3,923 | 47.94 |
| 2 | 1,599 | 19.54 |
| 3 | 915 | 11.18 |
| 4 | 601 | 7.34 |
| 5 | 372 | 4.55 |
| 6 | 259 | 3.17 |
| 7 | 185 | 2.26 |
| 8 | 87 | 1.06 |
| 9 | 153 | 1.87 |
| 10 or more | 89 | 1.09 |

Results of the full regression analysis are given in Appendix Tables 7 and 8. Female patients were more likely to be referred (OR 1.23; 95% CI 1.17-1.29) and increasing age had increasing odds of referral (OR 1.03; 95% CI 1.03-1.03, see Appendix Tables 5 and 6 for categorical results). The odds of being referred varied across WIMD quintiles with those in the least deprived quintile having the greatest odds of being referred (OR 1.39; 95% CI 1.29-1.48). Referrals were also more likely in rural locations (OR 1.17; 95% CI 1.12-1.22). There was no evidence of confounding within the multivariable regression modelling (Appendix tables 8 and 9).

## Discussion

Over the study period there was a large increase in patients seeking care from their GMP for persistent OFP. Attendances for all diagnoses increased, with the most pronounced increase in those attending with migraine and TMD. Patients tended to be female, and almost one-third attended more than once. Despite the number of attendances only a small proportion of patients were recorded as being referred for their persistent OFP. Factors associated with being referred included being female, increasing age and patient location.

As previously reported, limitations to this study include findings heavily relying on accurate Read code reporting by GMPs (Currie et al. 2022). Given the evidence suggesting that GMPs find persistent OFP difficult to diagnose there may be diagnostic errors which have translated into coding errors. An example of this could be age breakdown for BMS (Appendix Table 3) with a larger than expected number of young patients diagnosed with this. Additionally, some diagnoses may be underestimated as, for example, migraine can be misdiagnosed as sinusitis and TMD as otalgia (Kuttila et al. 2001). For this reason, the data by diagnosis should be interpreted with caution and rather the data presented is best regarded as a representation of overall burden of all persistent OFP diagnoses in primary medical care. The attendance rates for migraine are lower than previously reported (Latinovic et al. 2005), this may also be explained by coding inaccuracies, or all headache diagnoses being included in the reported study. In addition, within the Read codes available it was not possible to break diagnoses into acute and persistent diagnoses, therefore some patients who only attended once may actually represent an acute OFP presentation (for example acute TMD). However, given that OFP being present for more than 3 months significantly increases the likelihood of care seeking (Macfarlane et al. 2003) this could equally represent someone with a persistent presentation seeking care from their GMP for the first time. A strength of this study is the large sample size that was available for analysis over a long time period, meaning that issues with statistical power, external validity and generalisability were not a concern.

The increase in attendance rates for persistent OFP could be explained by either an increase in awareness or better reporting of persistent OFP diagnoses by GMPs, increasing NHS dental costs or access issues driving patients to attend their GMP rather than GDP, or an increase in incidence of persistent OFP and migraine. Given that by the end of the study period the rate of non-specific dental Read codes was reducing, this could perhaps reflect an increase in GMPs confidence in diagnosing persistent OFP. There is also evidence, however, that the prevalence and “chronification” of TMD is increasing (Häggman-Henrikson et al. 2020), as well as attendance rates for migraine (Lyngberg et al. 2005), and this could therefore represent an increase in number of patients seeking care as result of this. This maintenance of patients seeking care from their GMP for persistent OFP is in stark contrast to those seeking care for acute dental pain, whereby attendances dramatically reduced from 2006 and onwards (Currie et al. 2022).

Patients tended to be female across all diagnoses, which is in keeping with the wider literature on persistent OFP and migraine (Macfarlane et al. 2002; Latinovic et al. 2005; Koopman et al. 2009; Häggman-Henrikson et al. 2020). Young adults accounted for the greatest proportions for diagnoses of migraine and TMD, and in contrast, patients with BMS and atypical facial pain presented at a later age. The age groups at presentation are again in keeping with the expected age range for these diagnoses (Macfarlane et al. 2002; Koopman et al. 2009; Häggman-Henrikson et al. 2020). The frequency of adolescents attending with TMD was consistent with reported international findings (Christidis et al. 2019). Adolescents also attended almost as frequently as young adults with TMD supporting the suggestion that development of TMD in adolescence indicates an underlying vulnerability to musculoskeletal pain and increased likelihood of developing persistent pain from TMD into young adulthood (LeResche et al. 2007).

Patients presented from across all quintiles of WIMD with no obvious social gradient present. This is in contrast with patients presenting with acute dental pain, where there is a clear social gradient with patients from the most deprived areas being more likely to experience acute dental pain (Vargas et al. 2000; Steele et al. 2011; Currie, Stone, Brocklehurst, Slade, Durham, et al. 2022; Currie, Stone, Pearce, Landes, and Durham 2022). Chronic painful conditions, such as migraine (Burch et al. 2021), also tend to exhibit a social gradient, however there is mixed evidence within the persistent OFP literature (Von Korff et al. 1988; Andersson et al. 1993; Goulet et al. 1995; Aggarwal et al. 2003; Slade et al. 2013) and it is generally agreed that there is little association between persistent OFP and socioeconomic status. This finding therefore supports the existing literature showing lack of social gradient for persistent OFP, however is in contrast with existing literature on migraine. This could be explained by some the limitations of using WIMD in Wales where there are large rural areas where people from the most deprived areas tend to be more geographically dispersed and more disproportionately affected by some deprivation indictors (Jones 2015).

Most patients only attended their GMP once, however it is unknown whether they had also attended elsewhere, for example other primary care services such as their GDP, or secondary care services. Alternatively, these patients could reflect the 85% of patients with persistent pain who do not need treatment (Yekkalam and Wänman 2016), and as such were managed successfully by the GMP with education and self-management techniques and therefore did not require a further attendance. A proportion of patients attended on a repeated basis, with some patients having over ten attendances. This may reflect the complex care pathways these patients go through in an attempt to receive a diagnosis and manage their pain (Durham et al. 2011; Breckons et al. 2017). These repeated GMP attendances will be adding to the already established economic impact of persistent OFP (Durham et al. 2016), and may highlight the need to streamline care pathways in this patient group.

Despite the number of patients seeking care from their GMP only around 3% were referred suggesting that GMPs may feel comfortable managing these diagnoses in primary care, this is in contrast to a much higher rate of referrals from GMPs for acute dental pain presentations (Currie et al. 2022) for which GMPs are unable to treat. GMPs have previously reported feeling inadequately equipped to manage persistent OFP patients, however feel that they are obligated to treat them given they are able to manage patients with other long term chronic conditions (Peters et al. 2015), which could also explain the low number of referrals seen here. Alternatively, these patients may have been referred however an associated Read code not recorded, or, as previously reported (Breckons et al. 2017), they may have been informally referred to another service, such as their GDP. Regardless of the reasons for this, the low number of referrals seen here may not have the best outcome for patients with persistent OFP given that a failure to receive a diagnosis and appropriate management can lead to a worsening of symptoms (Durham et al. 2010; Durham et al. 2011; Breckons et al. 2017), as well as a breakdown in the doctor-patient relationship (Peters et al. 2015). This again suggests that care pathways for these patients can be improved, and that GMPs should perhaps be encouraged to refer persistent OFP patients at an earlier point to improve patient experience and outcome. Related to this, it is interesting to note the number of referrals in this study for suspected head and neck cancer. It is unknown whether these patients subsequently received a cancer diagnosis. However, if they did not, this care pathway is likely to have heightened patient concerns regarding diagnosis of their persistent pain and may subsequently have impacted negatively on their OFP outcome.

For patients who were referred, certain demographics were associated with a referral. Female patients were significantly more likely to be referred. This could be in keeping with the higher number of female patients experiencing persistent OFP, or the fact that female patients have a higher odds of being referred by GMPs for all conditions (Olthof et al. 2019). Patients presenting with persistent OFP diagnoses tend to be young adults or middle aged, which could explain why elderly patients had higher odds of being referred if they presented with new onset facial pain which could indicate a sinister underlying pathology. Adolescent patients presenting with OFP were the least likely age group to be referred despite the increased risk of “chronification” in these young patients (List et al. 2001). Patient location was also associated with receiving a referral, with patients in rural areas being more likely to be referred. The reasons for this are unknown, however, this could relate to dental access issues in rural areas for these patients if informal referrals between GMPs and GDPs are not possible, resulting in a referral being made at an earlier stage. Finally, patients from the most deprived areas were less likely to be referred, which given the equal spilt in presentations by WIMD could indicate inequalities in being referred for specialist management which may warrant further research.

In conclusion, an increasing number of patients are seeking care from their GMP for persistent OFP. These patients can attend on a repeated basis and very few are referred, however when they are referred this may occur multiple times. Predictors of receiving a referral for persistent OFP include female gender, older age and patient location.

## Data Availability

All data produced in the present work are contained in the manuscript

## Acknowledgements

CC Currie (Doctoral Research Fellow, NIHR DRF-2017-10-022) is funded by the National Institute for Health Research (NIHR) for this research project. The views expressed in this publication are those of the authors and not necessarily those of the NIHR, NHS or the UK Department of Health and Social Care. This study makes use of anonymised data held in the Secure Anonymised Information Linkage (SAIL) system, which is part of the national e-health records research infrastructure for Wales. We would like to acknowledge all the data providers who make anonymised data available for research. The responsibility for the interpretation of the information supplied by SAIL is the authors’ alone. The authors have no conflicts of interest to declare.

## Author Contributions

C Currie contributed to conception, design, data acquisition and interpretation, performed all statistical analyses, drafted and critically revised the manuscript. SJ Stone, P Brocklehurst, MS Pearce and J Durham contributed to conception, design, data acquisition and interpretation, and critically revised the manuscript. J Palmer contributed to data interpretation and drafted and critically revised the manuscript. VR Aggarwal and P Dorman contributed to data interpretation and critically revised the manuscript.

All authors gave their final approval and agree to be accountable for all aspects of the work.

## References

Aggarwal R, McBeth J, Zakrzewska JM, Lunt M, Macfarlane GJ. 2006. The epidemiology of chronic syndromes that are frequently unexplained: do they have common associated factors? Int J Epidemiol. 35(2):468–476. doi:10.1093/ije/dyi265.

Aggarwal VR, Macfarlane TV, Macfarlane GJ. 2003. Why is pain more common amongst people living in areas of low socio-economic status? A population-based cross-sectional study. Br Dent J. 194(7):383–387.

Anderson R, Richmond S, Thomas DW. 1999. Patient presentation at medical practices with dental problems: An analysis of the 1996 General Practice Morbidity Database for Wales. Br Dent J. 186(6):297–300.

Andersson HI, Ejlertsson G, Leden I, Rosenberg C. 1993. Chronic pain in a geographically defined general population: studies of differences in age, gender, social class and pain localization. Clin J Pain. 9:174–182.

Breckons M, Bissett SM, Exley C, Araujo-Soares V, Durham J. 2017. Care Pathways in Persistent Orofacial Pain: Qualitative Evidence from the DEEP Study. JDR Clin Transl Res. 2(1):48–57. doi:10.1177/2380084416679648.

Breckons M, Shen J, Bunga J, Vale L, Durham J. 2018. DEEP Study: Indirect and Out-of-pocket Costs of Persistent Orofacial Pain. J Dent Res. 97(11):1200–1206. doi:10.1177/0022034518773310.

Breivik H, Collett B, Ventafridda V, Cohen R, Gallacher D. 2006. Survey of chronic pain in Europe: prevalence, impact on daily life, and treatment. Eur J Pain. 10:287–333.

Burch R, Rizzoli P, Loder E. 2021. The prevalence and impact of migraine and severe headache in the United States: Updated age, sex, and socioeconomicJspecific estimates from government health surveys. Headache J Head Face Pain. 61(1):60–68. doi:10.1111/head.14024.

Christidis N, Lindström Ndanshau E, Sandberg A, Tsilingaridis G. 2019. Prevalence and treatment strategies regarding temporomandibular disordersin children and adolescents-A systematic review. J Oral Rehabil. 46:291–301.

Cope AL, Chestnutt IG, Wood F, Francis NA. 2016. Dental consultations in UK general practice and antibiotic prescribing rates: a retrospective cohort study. Br J Gen Pract. 66(646):e329–36. doi:10.3399/bjgp16X684757.

Currie CC, Stone SJ, Brocklehurst P, Slade G, Durham J, Pearce MS. 2022. Dental Attendances to General Medical Practitioners in Wales: A 44 Year-Analysis. J Dent Res. 101(4):407–413. doi:10.1177/00220345211044108.

Currie CC, Stone SJ, Pearce M, Landes D, Durham J. 2022. Urgent dental care use in the North East and Cumbria: predicting repeat attendance. Br Dent J. 232(3):164–171. doi:10.1038/s41415-022-3886-6.

Durham J, Breckons M, Vale L, Shen J. 2021. DEEP Study: Modeling Outcomes and Costs of Persistent Orofacial Pain. JDR Clin Transl Res.:238008442110638. doi:10.1177/23800844211063870.

Durham J, Shen J, Breckons M, Steele JG, Araujo-Soares V, Exley C, Vale L. 2016. Healthcare Cost and Impact of Persistent Orofacial Pain: The DEEP Study Cohort. J Dent Res. 95(10):1147–1154. doi:10.1177/0022034516648088.

Durham J, Steele J, Moufti MA, Wassell R, Robinson P, Exley C. 2011. Temporomandibular disorder patients’ journey through care: TMD patients’ journey through care. Community Dent Oral Epidemiol. 39(6):532–541. doi:10.1111/j.1600-0528.2011.00608.x.

Durham J, Steele JG, Wassell RW, Exley C. 2010. Living with Uncertainty: Temporomandibular Disorders. J Dent Res. 89(8):827–830. doi:10.1177/0022034510368648.

Ford DV, Jones KH, Verplancke JP, Lyons RA, John G, Brown G, Brooks CJ, Thompson S, Bodger O, Couch T, et al. 2009. The SAIL Databank: building a national architecture for e-health research and evaluation. BMC Health Serv Res. 9(1):157. doi:10.1186/1472-6963-9-157.

Goulet JP, Lavigne GJ, Lund JP. 1995. Jaw pain prevalence among French-speaking Canadians in Quebec and related symptoms of temporomandibular disorders. J Dent Res. 74:1738–1744.

Häggman-Henrikson B, Liv P, Ilgunas A, Visscher CM, Lobbezoo F, Durham J, Lövgren A. 2020. Increasing gender differences in the prevalence and chronification of orofacial pain in the population. Pain. 161(8):1768–1775. doi:10.1097/j.pain.0000000000001872.

Headache Classification Subcommitee of the International Headache Society. 2013. The international classification of headache disorders 3rd edition. Cephalgia. 33(9):629–808.

International Classification of Orofacial Pain. 2020. International Classification of Orofacial Pain, 1st edition (ICOP). Cephalalgia. 40(2):129–221. doi:10.1177/0333102419893823.

Jones L. 2015. Welsh Index of Multiple Deprivation 2014: A guide to analysing deprivation in rural areas. Welsh Gov. [accessed 2021 Apr 14]. https://gov.wales/sites/default/files/statistics-and-research/2019-05/welsh-index-of-multiple-deprivation-2014-a-guide-to-analysing-deprivation-in-rural-areas.pdf.

Koopman JSHA, Dieleman JP, Huygen FJ, de Mos M, Martin CGM, Sturkenboom MCJM. 2009. Incidence of facial pain in the general population. Pain. 147(1):122–127. doi:10.1016/j.pain.2009.08.023.

Kuttila SJ, Kuttila MH, Niemi PM, Le Bell YB, Alanen PJ, Suonpaa JT. 2001. Secondary otalgia in an adult population. Arch Otolaryngol Head Neck Surg. 127:401–405.

Latinovic R, Gulliford M, Ridsdale L. 2005. Headache and migraine in primary care: consultation, prescription, and referral rates in a large population. J Neurol Neurosurg Psychiatry. 77(3):385–387. doi:10.1136/jnnp.2005.073221.

LeResche L, Mancl LA, Drangsholt MT, Huang G, Von Korff M. 2007. Predictors of onset of facial pain and temporomandibular disorders in early adolescence. Pain. 129:269–278.

List T, Wahlund K, Larsson B. 2001. Psychosocial Functioning and Dental Factors in Adolescents with Temporomandibular Disorders: A Case-Control Study. J Orofac Pain. 15:218–27.

Lyngberg AC, Rasmussen BK, Jørgensen T, Jensen R. 2005. Secular Changes in Health Care Utilization and Work Absence for Migraine and Tension-type Headache: A Population Based Study. Eur J Epidemiol. 20(12):1007–1014. doi:10.1007/s10654-005-3778-5.

Macfarlane TV, Blinkhorn AS, Davies RM, Kincey J, Worthington HV. 2002. Oro-facial pain in the community: prevalence and associated impact. Community Dent Oral Epidemiol. 30:52–60.

Macfarlane TV, Blinkhorn AS, Davies RM, Kincey J, Worthington HV. 2003. Factors associated with health care seeking behaviour for orofacial pain in the general population. Community Dent Health. 20:20–26.

Maixner W, Diatchenko L, Dubner R, Fillingim RB, Greenspan JD, Knott C, Ohrbach R, Weir B, Slade GD. 2011. Orofacial Pain Prospective Evaluation and Risk Assessment Study – The OPPERA Study. J Pain. 12(11):T4–T11.e2. doi:10.1016/j.jpain.2011.08.002.

Olthof M, Groenhof F, Berger MY. 2019. Continuity of care and referral rate: challenges for the future of health care. Fam Pract. 36(2):162–165. doi:10.1093/fampra/cmy048.

Peters S, Goldthorpe J, McElroy C, King E, Javidi H, Tickle M, Aggarwal VR. 2015. Managing chronic orofacial pain: A qualitative study of patients’, doctors’, and dentists’ experiences. Br J Health Psychol. 20(4):777–791. doi:10.1111/bjhp.12141.

Réus JC, Polmann H, Souza BDM, Flores-Mir C, Gonçalves DAG, de Queiroz LP, Okeson J, De Luca Canto G. 2022. Association between primary headaches and temporomandibular disorders. J Am Dent Assoc. 153(2):120–131.e6. doi:10.1016/j.adaj.2021.07.021.

Shueb SS, Nixdorf DR, John MT, Alonso BF, Durham J. 2015. What is the impact of acute and chronic orofacial pain on quality of life? J Dent. 43(10):1203–1210. doi:http://dx.doi.org/10.1016/j.jdent.2015.06.001.

Slade GD, Bair E, Greenspan JD, Dubner R, Fillingim RB, Diatchenko L, Maixner W, Knott C, Ohrbach R. 2013. Signs and Symptoms of First-Onset TMD and Sociodemographic Predictors of Its Development: The OPPERA Prospective Cohort Study. J Pain. 14(12):T20–T32.e3. doi:10.1016/j.jpain.2013.07.014.

Steele JG, Pitts N, Fuller E, Treasure E. 2011. Urgent Conditions – a report from the Adult Dental Health Survey 2009. [accessed 2020 May 2]. https://files.digital.nhs.uk/publicationimport/pub01xxx/pub01086/adul-dent-heal-surv-summ-them-the3-2009-rep5.pdf.

Vargas CM, Macek MD, Marcus SE. 2000. Sociodemographic correlates of tooth pain among adults: United states, 1989. Pain. 85(1–2):87–92. doi:10.1016/s0304-3959(99)00250-x.

Von Korff M, Dworkin SF, Le Resche L, Kruger A. 1988. An epidemiologic comparison of pain complaints. Pain. 32:173–183. doi:10.1016/0304-3959(88)90066-8.

Yekkalam N, Wänman A. 2016. Factors associated with clinical decision-making in relation to treatment need for temporomandibular disorders. Acta Odontol Scand. 74(2):134–141. doi:10.3109/00016357.2015.1063159.

