## Supplemental Appendix for "Persistent Orofacial Pain Attendances at General Medical Practitioners"

**Supplemental Methodological Details**

**Welsh Index of Multiple Deprivation (WIMD)**

WIMD is the official measurement of deprivation of areas of Wales (Welsh Government, 2011) and takes into account eight different domains of deprivation: employment; income; education; health; community safety; geographical access to services; housing and physical environment. WIMD quintiles were used for analysis with quintile 1 being the 20% most deprived areas, and quintile 5 being the 20% least deprived areas of Wales.

**Office for National Statistics Urban/Rural Classification 2001**

The Office for National Statistics Urban/Rural classification 2001 (Office for National Statistics, 2016) divides geographical areas in urban (physical settlements with a population more than 10,000) and rural categories, with further subdivisions by settlement type and sparsity. Settlements in sparse areas have a particularly low number of households (compared to those defined as less sparse) and therefore may have implications on service availability.

**Calculation of Attendance Rates**

The attendance rate was calculated as the annual rate of dental consultations per 1000 patient-years to allow direct comparison with previous studies. The denominator to calculate the attendance rates was calculated using data from the Welsh Demographic Service (WDS) dataset available through SAIL. This patient level dataset contains all primary care events coded with Read codes by GMPs. This allowed calculation of the total patient-years for all attendances at all GMPs registered with SAIL by year. For partial annual data, e.g. where a patient entered the dataset part way through a calendar year, the amount of time they had contributed to that year was included by calculation of the proportion. For example, where a patient had entered the dataset in the 7th month of the year they were considered to have contributed 0.5 patient-years (6 months). Data were not available from the WDS dataset on patient location or age, therefore incidence rates could not be calculated for these variables.

**Supplemental Appendix Tables**

**Appendix Table 1:** Read codes used for data extraction and re-grouping of Read codes.

| Read Code | Read Code Description | Group |
| --- | --- | --- |
| F26 | Migraine | Migraine |
| F300 | Post herpetic trigeminal neuralgia | Post herpetic trigeminal neuralgia |
| F301z | Trigeminal neuralgia | Trigeminal neuralgia |
| F302 | Atypical facial pain | Atypical facial pain |
| J046 | Temporomandibular disorder | Temporomandibular disorders (TMDs) |
| J0462 | TMJ internal derangement |  |
| J0464 | TMJ dysfunction pain syndrome |  |
| J0463 | Temporomandibular click |  |
| S5y11 | TMJ sprain |  |
| J08zC | Burning mouth syndrome | Burning mouth syndrome (BMS) |
| J096 | Burning tongue |  |
| SJ12 | Trigeminal (5^th^) nerve injury | Trigeminal nerve injury |
| J080 | Stomatitis | Stomatitis |
| 191.. | Tooth symptoms | Non-specific dental Read codes |
| J05y | Other specified dental disorder |  |
| J052 | Dental diseases/conditions |  |

**Appendix Table 2:** Detailed patient demographics from the entire dataset (1974-2017).

| **Patient Gender** | **All Diagnoses, n (%)** | **Excluding Migraine, n (%)** |
| --- | --- | --- |
| Male | 132,683 (23.34) | 59,969 (35.36) |
| Female | 335,454 (71.66) | 109,616 (64.64) |
| Indeterminate/anticipated sex change | 0 | 0 |
| Not known | 0 | 0 |
| **Patient Age Group** | **All Diagnoses, n (%)** | **Excluding Migraine, n (%)** |
| <10 years | 13,128 (2.80) | 5,897 (3.48) |
| 10-19 years | 66,334 (14.17) | 14,880 (8.77) |
| 20-29 years | 96,333 (20.58) | 29,964 (17.67) |
| 30-39 years | 87,264 (18.64) | 25,910 (15.28) |
| 40-49 years | 82,319 (17.58) | 26,408 (15.57) |
| 50-59 years | 58,312 (12.46) | 24,694 (14.56) |
| 60-69 years | 35,770 (7.64) | 20,712 (12.21) |
| 70-79 years | 20,097 (4.29) | 14,227 (8.39) |
| >80 years | 8,580 (1.83) | 6,893 (4.06) |
| **WIMD Quintile, n (%)** | **All Diagnoses, n (%)** | **Excluding Migraine, n (%)** |
| 1 (most deprived) | 96,285 (20.57) | 34,674 (20.45) |
| 2 | 89,722 (19.17) | 32,728 (19.30) |
| 3 | 96,167 (20.54) | 36,154 (21.32) |
| 4 | 87,777 (18.75) | 32,899 (19.40) |
| 5 (least deprived) | 98,186 (20.97) | 33,130 (19.54) |
| **Urban/Rural Definition, n (%)** | **All Diagnoses, n (%)** | **Excluding Migraine, n (%)** |
| Urban; sparse | 14,652 (3.13) | 6,104 (3.6) |
| Urban; less sparse | 293,928 (62.79) | 100,968 (59.54) |
| Town & fringe; sparse | 16,463 (3.52) | 7,567 (4.46) |
| Town & fringe; less sparse | 67,333 (14.38) | 24,571 (14.49) |
| Village, hamlet & isolated dwellings; sparse | 41,394 (3.13) | 17,669 (10.42) |
| Village, hamlet & isolated dwellings; less sparse | 34,367 (7.34) | 12,706 (7.49) |

**Appendix Table 3**: Detailed patient demographics by diagnosis over the entire dataset (1974-2017). **Note: counts <5 were present in subgroup analysis therefore breakdown is only shown for the most common diagnoses to protect patient confidentiality.**

|  | **Migraine** | | **TMD** | | **BMS** | | **Atypical facial pain** | |
| --- | --- | --- | --- | --- | --- | --- | --- | --- |
|  | **n** | **%** | **n** | **%** | **n** | **%** | **n** | **%** |
| **Gender** | | | | | | | |  |
| Male | 72,714 | 24.36 | 16,291 | 28.29 | 2,462 | 29.69 | 2,022 | 27.39 |
| Female | 225,838 | 75.64 | 41,509 | 71.81 | 5,829 | 70.31 | 5,361 | 72.61 |
| **WIMD** | | | | | | | |  |
| 1 | 61,611 | 20.64 | 10,764 | 18.62 | 1,571 | 18.95 | 1,288 | 17.45 |
| 2 | 56,994 | 19.09 | 10,849 | 18.77 | 1,442 | 17.39 | 1,434 | 19.42 |
| 3 | 60,013 | 20.10 | 11,552 | 19.99 | 1,736 | 20.94 | 1,554 | 21.05 |
| 4 | 54,878 | 18.38 | 10,990 | 19.01 | 1,623 | 19.58 | 1,431 | 19.38 |
| 5 | 65,056 | 21.79 | 13,645 | 23.61 | 1,919 | 23.15 | 1,676 | 22.70 |
| **Age Group** | | | | | | | |  |
| <10 | 7,231 | 2.42 | 175 | 0.30 | 533 | 6.43 | 104 | 1.41 |
| 10-19 | 51,454 | 17.23 | 8,221 | 14.22 | 421 | 5.08 | 265 | 3.59 |
| 20-29 | 66,369 | 22.23 | 11,001 | 19.03 | 601 | 7.25 | 695 | 9.41 |
| 30-39 | 61,354 | 20.55 | 9,293 | 16.08 | 775 | 9.35 | 1,113 | 15.07 |
| 40-49 | 55,911 | 18.72 | 8,972 | 15.52 | 1,099 | 13.26 | 1,451 | 19.65 |
| 50-59 | 33,618 | 11.26 | 8,147 | 14.10 | 1,351 | 16.29 | 1,457 | 19.73 |
| 60-69 | 15,058 | 5.04 | 6,547 | 11.33 | 1,604 | 19.34 | 1,164 | 15.77 |
| 70-79 | 5,870 | 1.97 | 4,007 | 6.93 | 1,259 | 15.19 | 740 | 10.02 |
| >80 | 1,687 | 0.57 | 1,437 | 2.49 | 648 | 7.82 | 394 | 5.34 |
| **Total** | 298,552 | 100.00 | 57,800 | 100.00 | 8,291 | 100.00 | 7,383 | 100.00 |

**Appendix Table 4:** Number of times patients with a diagnosis of both TMD and migraine attended over the time period studied.

| **Number of Attendances** | **Number** | **%** |
| --- | --- | --- |
| **1** | 0 | 0 |
| **2** | 1,187 | 21.55 |
| **3** | 1,136 | 20.62 |
| **4** | 821 | 14.91 |
| **5** | 570 | 10.35 |
| **6** | 437 | 7.93 |
| **7** | 264 | 4.79 |
| **8** | 234 | 4.25 |
| **9** | 163 | 2.96 |
| **10 or more** | 696 | 12.64 |
| **Total** | 5,508 | 100.00 |

**Appendix Table 5:** Breakdown of referral Read codes associated with a chronic OFP diagnosis. *NB dental referrals have been regrouped to due to counts of <5. Dental referrals were made to the following specialties: restorative dentistry; paediatric dentistry; oral surgery; endodontics; dental radiology; non-specific dental services referral.

| **Referral Code** | **Number** | **%** |
| --- | --- | --- |
| Referral needed | 8,710 | 43.33 |
| Further care referral NOS | 3,441 | 17.12 |
| Ref to clinic | 2,900 | 14.43 |
| Ref to dental services* | 1,576 | 7.84 |
| Ref to other care | 867 | 4.31 |
| Referred to service | 590 | 2.93 |
| Ref to other clinic | 557 | 2.77 |
| Fast track suspected H&N cancer | 448 | 2.23 |
| Ref to clinic NOS | 334 | 1.66 |
| Private ref to maxfax | 273 | 1.36 |
| Private ref to oral surgeon | 170 | 0.85 |
| Informal referral, signposted to other service | 129 | 0.64 |
| Private ref to pain management | 108 | 0.54 |
| Total | 20,103 | 100.00 |

**Appendix Table 6:** Number of referrals associated with diagnostic Read codes. NB data on referrals for post-herpetic trigeminal neuralgia and trigeminal nerve injury are excluded due to counts present <5. *indicates counts regrouped to overcome counts <5 in atypical facial pain.

| **Number of Referrals** | **Migraine** | **TMD** | **BMS** | **Atypical Facial Pain** | **Trigeminal Neuralgia** | **Non-specific dental** |
| --- | --- | --- | --- | --- | --- | --- |
| **1** | 2,848 | 772 | 115 | 77 | 243 | 2,006 |
| **2** | 577 | 140 | 25 | 14 | 40 | 322 |
| **3** | 219 | 43 | 9 | 8* | 14 | 109 |
| **4** | 115 | 20 | 5 | 0* | 8 | 49 |
| **5 or more** | 126 | 21 | 7 | 0* | 9 | 73 |

**Appendix Table 7:** Number of times patients with a diagnosis of both TMD and migraine were referred over the time period studied.

| **Number of Referrals** | **n** | **%** |
| --- | --- | --- |
| **0** | 5,245 | 95.23 |
| **1** | 208 | 3.78 |
| **2** | 28 | 0.51 |
| **3** | 11 | 0.20 |
| **4** | 7 | 0.13 |
| **5 or more** | 9 | 0.16 |
| **Total** | 5,508 | 100.00 |

**Appendix Table 8:** Logistic regression for referrals (all diagnoses) with adjustments in multivariable model.

|  | **Univariate Analysis** | | | **Adjusted for Age** | | | **Adjusted for Urban/Rural** | | | **Adjusted for Gender** | | | **Adjusted for WIMD** | | |
| --- | --- | --- | --- | --- | --- | --- | --- | --- | --- | --- | --- | --- | --- | --- | --- |
|  | **OR** | **95% CI** | **P Value** | **OR** | **95% CI** | **P Value** | **OR** | **95% CI** | **P Value** | **OR** | **95% CI** | **P Value** | **OR** | **95% CI** | **P Value** |
| **WIMD** | | | | | | | | | | | | | | | |
| 1 | 1.00 (ref) |  |  |  |  |  |  |  |  |  |  |  |  |  |  |
| 2 | 1.34 | 1.25-1.44 | <0.0001 | 1.30 | 1.21-1.39 | <0.0001 | 1.31 | 1,22-1.41 | <0.0001 | 1.34 | 1.25-1.44 | <0.0001 |  |  |  |
| 3 | 1.28 | 1.20-1.38 | <0.0001 | 1.18 | 1.10-1.27 | <0.0001 | 1.22 | 1.13-1.31 | <0.0001 | 1.28 | 1.20-1.38 | <0.0001 |  |  |  |
| 4 | 1.05 | 0.97-1.13 | 0.206 | 0.95 | 0.88-1.02 | 0.151 | 0.99 | 0.92-1.07 | 0.864 | 1.05 | 0.97-1.13 | 0.225 |  |  |  |
| 5 | 1.39 | 1.29-1.48 | <0.0001 | 1.21 | 1.13-1.30 | <0.0001 | 1.34 | 1.25-1.44 | <0.0001 | 1.38 | 1.28-1.48 | <0.0001 |  |  |  |
| **Urban/Rural** | | | | | | | | | | | | | | | |
| Urban | 1.00 (ref) |  |  |  |  |  |  |  |  |  |  |  |  |  |  |
| Rural | 1.17 | 1.12-1.22 | <0.0001 | 1.09 | 1.04-1.14 | <0.0001 |  |  |  | 1.17 | 1.12-1.22 | <0.0001 | 1.16 | 1.11-1.22 | <0.0001 |
| **Gender** | | | | | | | | | | | | | | | |
| Male | 1.00 (ref) |  |  |  |  |  |  |  |  |  |  |  |  |  |  |
| Female | 1.23 | 1.17-1.29 | <0.0001 | 1.22 | 1.16-1.28 | <0.0001 | 1.23 | 1.17-1.29 | <0.0001 |  |  |  | 1.22 | 1.16-1.29 | <0.0001 |
| **Age Group** | | | | | | | | | | | | | | | |
| <10 | 1.00 (ref) |  |  |  |  |  |  |  |  |  |  |  |  |  |  |
| 10-19 | 0.87 | 0.70-1.09 | 0.220 |  |  |  | 0.87 | 0.70-1.09 | 0.222 | 0.84 | 0.67-1.05 | 0.125 | 0.87 | 0.70-1.09 | 0.215 |
| 20-29 | 1.50 | 1.22-1.86 | <0.0001 |  |  |  | 1.51 | 1.22-1.86 | <0.0001 | 1.43 | 1.16-1.76 | <0.001 | 1.50 | 1.22-1.85 | <0.0001 |
| 30-39 | 1.99 | 1.62-2.45 | <0.0001 |  |  |  | 1.99 | 1.61-2.45 | <0.0001 | 1.89 | 1.53-2.33 | <0.0001 | 1.99 | 1.61-2.45 | <0.0001 |
| 40-49 | 2.45 | 1.99-3.01 | <0.0001 |  |  |  | 2.44 | 1.98-3.00 | <0.0001 | 2.32 | 1.88-2.86 | <0.0001 | 2.44 | 1.98-3.00 | <0.0001 |
| 50-59 | 3.43 | 2.79-4.22 | <0.0001 |  |  |  | 3.41 | 2.77-4.20 | <0.0001 | 3.26 | 2.65-4.02 | <0.0001 | 3.41 | 2.77-4.20 | <0.0001 |
| 60-69 | 4.44 | 3.60-5.47 | <0.0001 |  |  |  | 4.41 | 3.58-5.44 | <0.0001 | 4.27 | 3.47-5.27 | <0.0001 | 4.43 | 3.59-5.46 | <0.0001 |
| 70-79 | 5.34 | 4.31-6.60 | <0.0001 |  |  |  | 5.31 | 4.29-6.56 | <0.0001 | 5.14 | 4.15-6.36 | <0.0001 | 5.30 | 4.29-6.56 | <0.0001 |
| >80 | 6.89 | 5.52-8.60 | <0.0001 |  |  |  | 6.85 | 5.48-8.55 | <0.0001 | 6.59 | 5.27-8.22 | <0.0001 | 6.84 | 5.48-8.54 | <0.0001 |

**Appendix Table 9:** Logistic regression for referrals (excluding migraine) with adjustments in multivariable model.

|  | **Univariate Analysis** | | | **Adjusted for Age** | | | **Adjusted for Urban/Rural** | | | **Adjusted for Gender** | | | **Adjusted for WIMD** | | |
| --- | --- | --- | --- | --- | --- | --- | --- | --- | --- | --- | --- | --- | --- | --- | --- |
|  | **OR** | **95% CI** | **P Value** | **OR** | **95% CI** | **P Value** | **OR** | **95% CI** | **P Value** | **OR** | **95% CI** | **P Value** | **OR** | **95% CI** | **P Value** |
| **WIMD** | | | | | | | | | | | | | | | |
| 1 | 1.00 (ref) |  |  |  |  |  |  |  |  |  |  |  |  |  |  |
| 2 | 1.33 | 1.21-1.50 | <0.05 | 1.31 | 1.19-1.45 | <0.0001 | 1.30 | 1.18-1.44 | <0.0001 | 1.33 | 1.20-1.46 | <0.0001 |  |  |  |
| 3 | 1.18 | 1.07-1.31 | <0.001 | 1.14 | 1.03-1.26 | <0.001 | 1.12 | 1.01-1.24 | 0.032 | 1.77 | 1.07-1.30 | <0.01 |  |  |  |
| 4 | 0.96 | 0.86-1.07 | 0.443 | 0.92 | 0.83-1.02 | 0.123 | 0.90 | 0.81-1.01 | 0.073 | 0.95 | 0.86-1.06 | <0.362 |  |  |  |
| 5 | 1.36 | 1.24-1.50 | <0.0001 | 1.28 | 1.17-1.42 | <0.0001 | 1.32 | 1.20-1.46 | <0.0001 | 1.35 | 1.22-1.48 | <0.0001 |  |  |  |
| **Urban/Rural** | | | | | | | | | | | | | | | |
| Urban | 1.00 (ref) |  |  |  |  |  |  |  |  |  |  |  |  |  |  |
| Rural | 1.13 | 1.06-1.20 | <0.0001 | 1.10 | 1.04-1.17 | <0.01 |  |  |  | 1.13 | 1.06-1.20 | <0.0001 | 1.15 | 1.08-1.23 | <0.0001 |
| **Gender** | | | | | | | | | | | | | | | |
| Male | 1.00 (ref) |  |  |  |  |  |  |  |  |  |  |  |  |  |  |
| Female | 1.49 | 1.39-1.60 | <0.0001 | 1.47 | 1.38-1.58 | <0.0001 | 1.49 | 1.39-1.60 | <0.0001 |  |  |  | 1.49 | 1.39-1.59 | <0.0001 |
| **Age Group** | | | | | | | | | | | | | | | |
| <10 | 1.00 (ref) |  |  |  |  |  |  |  |  |  |  |  |  |  |  |
| 10-19 | 1.23 | 0.97-1.55 | 0.082 |  |  |  | 1.23 | 0.97-1.55 | 0.082 | 1.15 | 0.91-1.46 | 0.237 | 1.22 | 0.97-1.55 | 0.091 |
| 20-29 | 1.31 | 1.06-1.63 | 0.014 |  |  |  | 1.32 | 1.06-1.64 | <0.014 | 1.25 | 1.00-1.55 | <0.05 | 1.31 | 1.05-1.63 | <0.05 |
| 30-39 | 1.72 | 1.39-2.14 | <0.0001 |  |  |  | 1.72 | 1.34-2.14 | <0.0001 | 1.64 | 1.32-2.03 | <0.0001 | 1.71 | 1.38-2.13 | <0.0001 |
| 40-49 | 1.75 | 1.41-2.17 | <0.0001 |  |  |  | 1.74 | 1.40-2.16 | <0.0001 | 1.65 | 1.33-2.04 | <0.0001 | 1.74 | 1.40-2.15 | <0.0001 |
| 50-59 | 1.79 | 1.45-2.23 | <0.0001 |  |  |  | 1.78 | 1.44-2.21 | <0.0001 | 1.69 | 1.36-2.10 | <0.0001 | 1.78 | 1.43-2.21 | <0.0001 |
| 60-69 | 1.87 | 1.51-2.33 | <0.0001 |  |  |  | 1.86 | 1.50-2.31 | <0.0001 | 1.78 | 1.43-2.21 | <0.0001 | 1.86 | 1.49-2.31 | <0.0001 |
| 70-79 | 2.07 | 1.65-2.58 | <0.0001 |  |  |  | 2.05 | 1.65-2.56 | <0.0001 | 1.96 | 1.57-2.44 | <0.0001 | 2.04 | 1.63-2.54 | <0.0001 |
| >80 | 2.68 | 2.13-3.39 | <0.0001 |  |  |  | 2.67 | 2.12-3.37 | <0.0001 | 2.49 | 1.97-3.14 | <0.0001 | 2.65 | 2.10-3.34 | <0.0001 |
